# High-gamma electrocorticography activity represents perceived vibration intensity in human somatosensory cortex

**DOI:** 10.1101/2025.07.09.25331186

**Authors:** Oranatt Chaichanasittikarn, Lauren Diaz, Neha Thomas, Daniel Candrea, Shiyu Luo, Kevin Nathan, Francesco V. Tenore, Matthew S. Fifer, Nathan E. Crone, Breanne Christie, Luke E. Osborn

## Abstract

Haptic feedback can play a useful role in rehabilitation and brain-computer interface applications by providing users with information about their system or performance. One challenge delivering tactile stimulation is not knowing how the haptic sensation is actually perceived, irrespective of the stimulation amplitude, during real-world use and beyond controlled psychophysical experiments. In a participant with chronically implanted electrocorticography arrays, we observed that perceived intensity of haptic vibration on the fingertips was represented in the high-gamma (HG) frequency band (70-170 Hz) in the somatosensory cortex. The five fingers of the participant’s right hand were represented by distinct channels in the implanted array and modulated by the vibration amplitude at the fingertips. Although it reliably varied with the vibration amplitude, we found that HG activity had a stronger relationship with the actual perceived intensity of haptic stimulation (*r_s_*= 0.45*, p <* 10^−6^). These results demonstrate that neural signals, specifically HG activity, in the somatosensory cortex can represent qualities of perceived haptic intensity regardless of the stimulation amplitude, which could enable a new way to passively quantify or ensure effective haptic feedback to a user.

## I. Introduction

Haptic feedback enables information transfer to a user and can often lead to improved function or performance. Alongside advances in wearable haptic devices, touch feedback continues to be an important feature in human-centered systems [1]. Tactile sensations can enable greater prosthetic arm function [2] or help increase performance during vision-deficient tele-manipulation [3]. In the case of intracortical brain-computer interfaces (BCI), haptic sensations can be also elicited through electrical stimulation of the brain to convey object [4] or grip force [5] information during grasping. In another example, BCI cursor control was improved by using haptic stimulation of the skin [6].

Because of the value that haptic perception provides as a feedback signal to improve function, it is important that human-in-the-loop systems and any haptic feedback are engineered in a way that ensures reliable tactile perceptions. While traditional sensory characterization relies on psychophysical experiments [7], one possibility for monitoring tactile perception in real-time with BCI systems is by measuring relevant cortical activity.

Electrocorticography (ECoG) has been used for mapping finger representations in the somatosensory cortex [8]–[and prior work has suggested that touch intensity is also encoded in the same cortical region [11]. While ECoG signals in the high-gamma (HG) frequency reflect vibrotactile inputs on the skin and are modulated based on attention [12], there is also evidence that signals in the alpha and beta frequency ranges are also modulated by vibration intensity [13]. The firing of neurons in the somatosensory cortex has also been shown to be responsive to changes in mechanical indentation depth and rate [14], which are presumably correlated with the perceived intensity of touch events.

HG neural activity is modulated by a range of tactile experiences. In the somatosensory cortex, HG signals vary with vibrotactile frequency [15] and are also known to be different during deep touch versus light or soft touch events [16]. Others have shown that HG signals from implanted ECoG arrays in the somatosensory cortex are primarily representative of somatosensory feedback during movement [17] and that, in a rat model, HG frequency signals can be separated further to differentiate contact rate and absolute force applied during tactile events [18].

It is clear that tactile events and stimulation characteristics are reflected in HG neural signals; however, a remaining question is whether these signals are more reflective of the physical stimulation properties or the perceptual experiences and perceived qualities of the stimulation. For example, neural signals can be used to classify different emotions [19] and to decode intended loudness of speech [20]. In the case of touch, the somatosensory cortex responds differently to active and passive touch [21] and the expectation of different touch inputs and experiences also affects the cortical response [22]. One question that remains is whether neural signals in the somatosensory cortex reflect touch perception itself or are only representative of the haptic stimulation input.

To better understand if a neural physiological response reflects perceived intensity, not just stimulation amplitude, of a touch event, we delivered fingertip vibrations to an individual with implanted ECoG arrays and measured the reported levels of perceived intensity for varying vibration amplitudes. We observed that HG activity was modulated by both vibration amplitude and reported perceived intensity, but that the neural response was more strongly modulated by the perceived haptic intensity.

## II. Methods

This study was performed under the CortiCom clinical trial (NCT03567213) and was reviewed and approved by the Johns Hopkins University Institutional Review Board (IRB) and by the Food and Drug Administration (FDA) under an investigational device exemption.

A male participant living with amyotrophic lateral sclerosis (ALS), which had led to upper limb weakness, was implanted with two 8 × 8 subdural ECoG electrode grids (PMT Corporation) implanted in the left hemisphere over speech and upper extremity sensorimotor representations (Fig. 1A) [23]–[25]. Each ECoG array contained platinum-iridium electrodes, which were 4 mm apart (center-to-center) and covered an area of 36.6 mm × 33.1 mm [23]–[25].

**Fig. 1.**
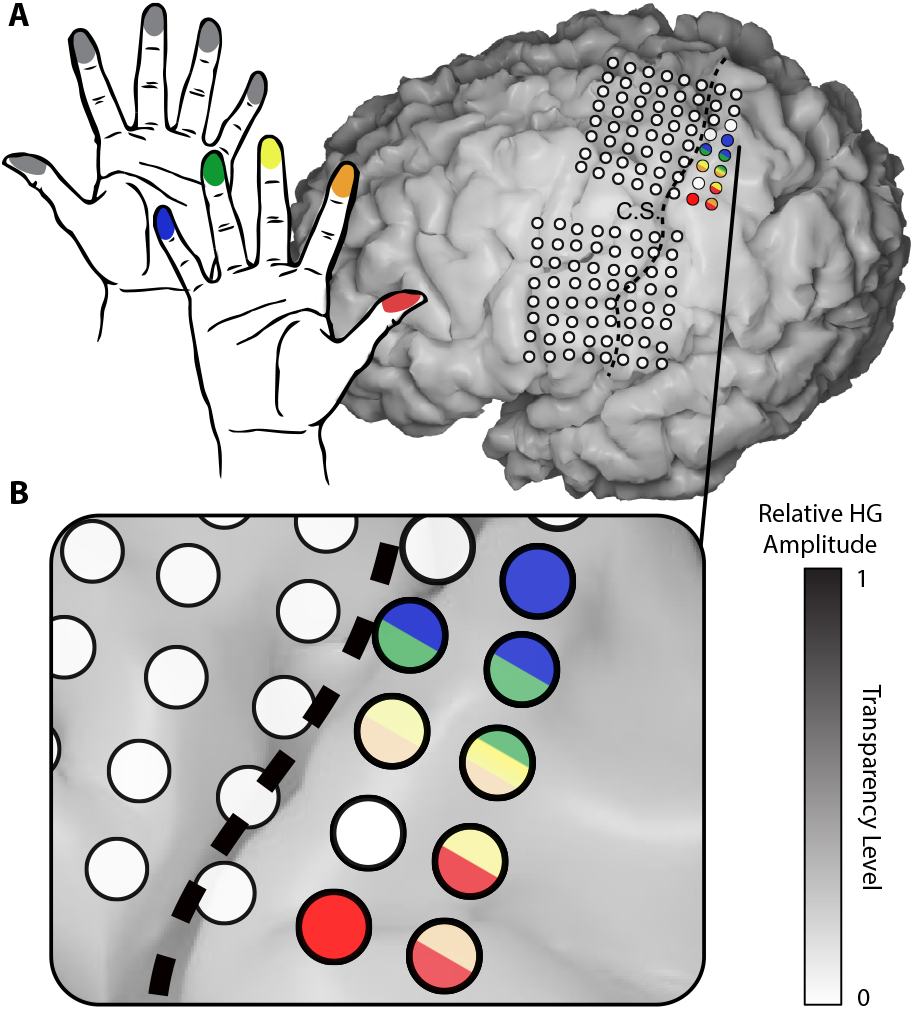
(A) Vibration was delivered to individual fingertips across both hands, but only vibration on the right hand fingers showed high HG activity (z-score *>* 2) in channels on the implanted arrays during haptic stimulation. The two ECoG arrays spanned the central sulcus (C.S.). All five fingers on the right hand were represented by channels covering the postcentral gyrus and the top three most responsive channels during haptic vibration are shown for each finger. (B) Zoomed in electrode array showing the top three channels for each finger. Color transparency indicates the relative HG activity z-score magnitude on each channel during stimulation.

### A. Experiment

A vibration tactor (C3, Engineering Acoustics, Inc) was placed on the tip of every finger on both the right and left hands. The participant was seated and a vibration stimulation was delivered to individual fingers using an amplitude of either 25% (low), 50% (medium), or 75% (high) of the maximum tactor amplitude. The vibration, which was 500 ms with a frequency of 300 Hz, was randomized in amplitude and location for each trial. The participant was instructed to attend to the vibration on his fingers. The C3 tactors were controlled using the Universal Controller (Engineering Acoustics, Inc) and have been used in our prior work to provide haptic vibration to human participants [26], [27].

On the right hand each vibration amplitude was presented 50 times on each finger for a total of 750 trials across all conditions. For each trial, the participant verbally reported the perceived intensity of the haptic vibration, using a scale from zero to ten with zero being no perceived haptic sensation and ten being the strongest sensation. Each experiment block contained 45 trials.

The participant also received vibration on the right and left hand fingertips but was instructed to not verbally report the perceived haptic intensities. Each vibration condition was presented 20 times on each finger for a total of 300 trials on the right hand and 300 trials on the left hand.

The ECoG electrodes were connected to a 128-ch Neuroport pedestal (Blackrock Neurotech) and sampled at 2 kHz. The ECoG signals were recorded during blocks of fingertip vibration during a single visit by the participant to the laboratory. A custom MATLAB script was used to control the vibration stimulation and BCI2000 [28] was used to control the data acquisition during each block.

### B. Data analysis

We used a Fast Fourier Transform (FFT) and computed the spectral power of 128 ms windows shifted by 64 ms increments for each of the 128 ECoG channels. The HG spectral power was calculated as the mean spectral power in the frequency band between 70 and 170 Hz.

For each channel, the HG spectral power for each trial was normalized by using the average signal between −500 ms to 100 ms, relative to vibration onset, as the baseline. To calculate the HG activity during haptic vibration events, we averaged the HG spectral power of the top five channels for each finger between 0 ms and 300 ms, relative to vibration onset. Signals from each trial were then grouped by vibration condition (i.e., location and amplitude) and averaged together.

To identify the mapping of the hand and to determine which ECoG channels were modulated by haptic vibration, we calculated the HG spectral power and set a threshold of zscore *>*2 to determine which channels responded to vibration on each finger. To estimate the effective vibration amplitude for each trial, we calculated the mean peak voltage of the vibration waveform.

For the regression models, the top five channels with the highest HG spectral power were selected for each finger under each vibration condition. The HG spectral power of each group was z-score standardized prior to analysis. A linear regression analysis was performed using the least squares method to assess the relationship between the HG spectral power and both the vibration amplitude and the perceived amplitude. Spearman correlation was performed to measure strength of monotonic relationship between the HG spectral power and both the vibration amplitude and the perceived amplitude.

Unless otherwise noted, statistical p-values were calculated using a two-sided Mann-Whitney *U* test. The whiskers on the violin plots represent the minimal and maximal values, the vertical lines indicate the first and third quartiles, and the white dots represent the median of the distribution. All analyses were performed with MATLAB and Python.

## III. Results &Discussion

### A. Finger representation

First, to identify hand representation in the implanted electrode arrays, we delivered haptic vibration using the “high” amplitude on the fingertips while measuring HG activity. Vibration on the five fingers on the right hand led to significant modulation (z-score *>* 2) for each finger in at least one unique channel covering the postcentral gyrus (Fig. 1). The right thumb and right little finger were most strongly represented with HG z-scores of 5.8 and 6.8, respectively, during the haptic stimulation.

None of the fingers on the left hand appeared to be strongly represented by HG activity in the implanted ECoG arrays. However, the left thumb did show slightly elevated HG activity (z-score 1.6-1.8) during haptic vibration events on the same three electrodes shown in Fig. 1 that responded to right thumb vibration. This observed, albeit weak, ipsilateral representation of the hand in the somatosensory cortex has previously been reported [29]; however, it was also suggested that ipsilateral finger representations are driven by active movement and not passive sensory inputs [29].

### B. High-gamma response to vibration amplitude

Modulation of HG activity was observed as a function of the amplitude of haptic vibration at the fingertips (Fig. 2). The HG z-score from single channels during thumb and little finger vibration is shown in Fig. 2A. When combining the top five channels for each vibration location, there was a significant difference (*p <* 0.05) in HG activity between all three vibration amplitudes (Fig. 2B). The median z-score during haptic stimulation was 2.6, 2.7, and 3.4 for the low, medium, and high stimulation amplitudes, respectively. Although individual z-score distributions are statistically distinct, the median value was most notably different for the high vibration amplitude condition.

**Fig. 2.**
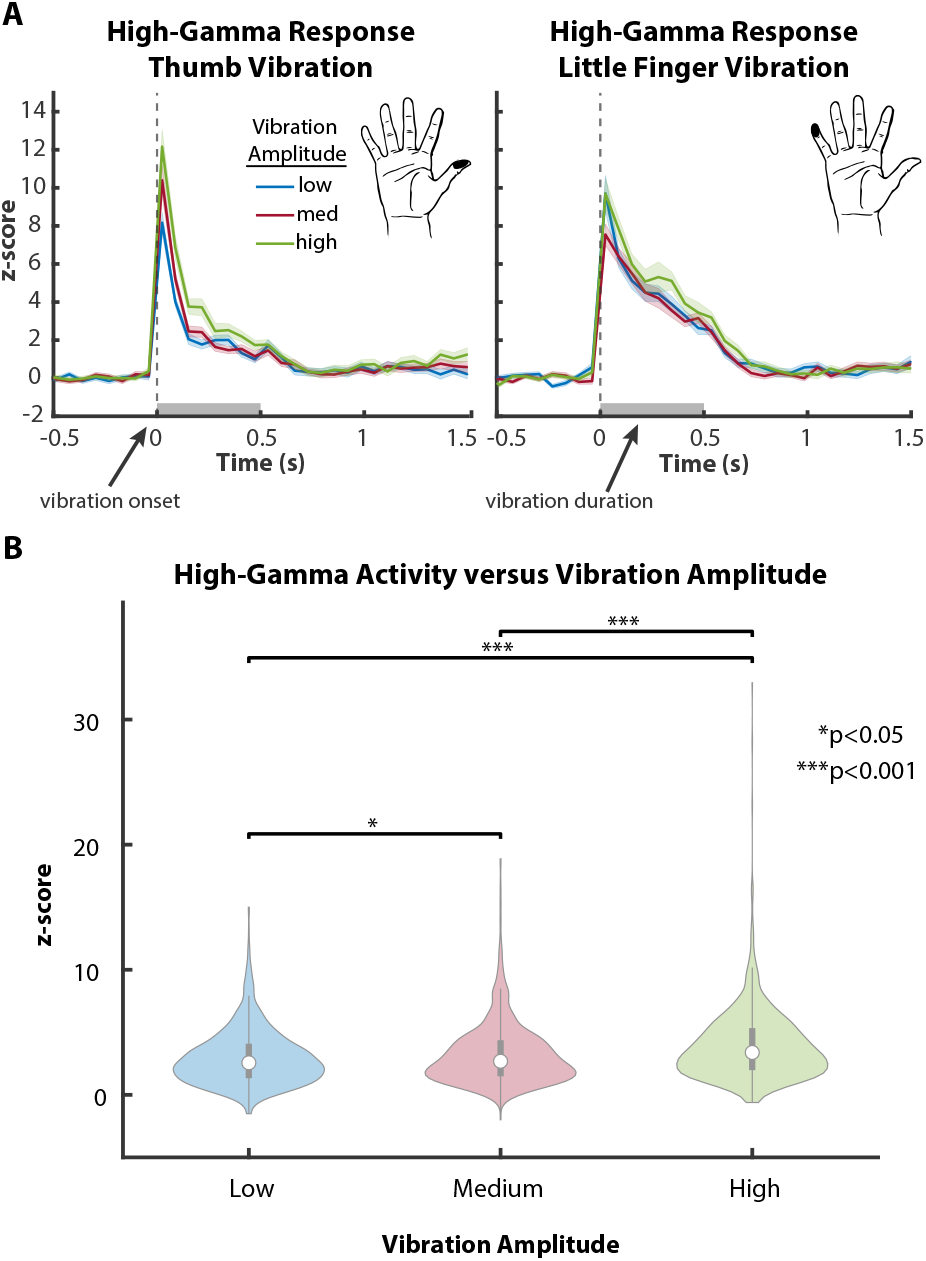
(A) Exemplary HG response from a single channel for low, medium, and high vibration amplitudes on the right thumb and little finger. Slight, but observable differences exist in the HG activity. (B) Average HG activity during haptic vibration events combined across the top five channels and all fingers on the right hand.

### C. High-gamma response to perceived intensity

To understand how the perceived intensity of haptic vibration was reflected in the neural activity, we next grouped trials by the reported intensity (Fig. 3). The participant perceived the vibration on every trial and his verbally reported intensities ranged from one to eight. When comparing HG z-score to the reported intensity levels, there is an increasing trend with larger HG amplitudes occurring on trials with a higher reported intensity. The median z-score during haptic stimulation was 1.8, 2.0, 3.0, and 4.3 for perceived intensities of one through four, respectively (Fig. 3). There were only two trials where the participant reported an intensity of five and only one trial each for intensities of seven and eight. The participant did not report any trials as a “six”.

**Fig. 3.**
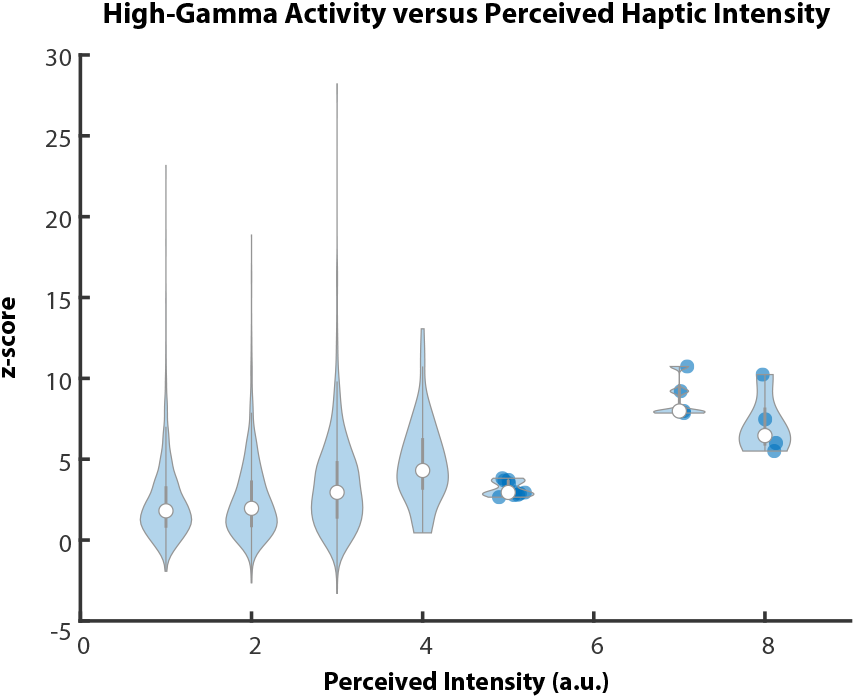
Average HG activity across the top five channels and all fingers on the right hand as a function of the reported perceived intensity. Data points are shown on the plot for reported intensities with fewer than 50 data points. Kruskal–Wallis *p <* 0.001.

Compared to HG modulation when grouping trials by vibration amplitude, there was larger z-score modulation and a wider range of perceived amplitudes. This difference in modulation suggests that the HG activity during haptic vibration may better reflect the actual experienced tactile sensation intensity compared to the physical amplitude of the mechanical indentations of the vibrating tactor.

Prior work demonstrated that attention can influence the HG representations of tactile events in human participants [12], so it is possible that changes in reported perceived intensity are also related to changes in attention across different trials, which leads to modulation of HG activity. Regardless, our results show that the HG amplitude during haptic vibration reflects the perceived intensity of the sensation.

### D. Relationship between perceived and actual vibration

We then explored how the different vibration amplitudes were subjectively rated by the participant. In order to make this comparison, we first verified the voltage of the tactor output to calculate an effective amplitude for each trial (Fig. 4A). The tactors had some variation in their output on each trial for a given amplitude condition, but the distributions of the average voltages were significantly different (*p <* 0.001) across all three conditions.

**Fig. 4.**
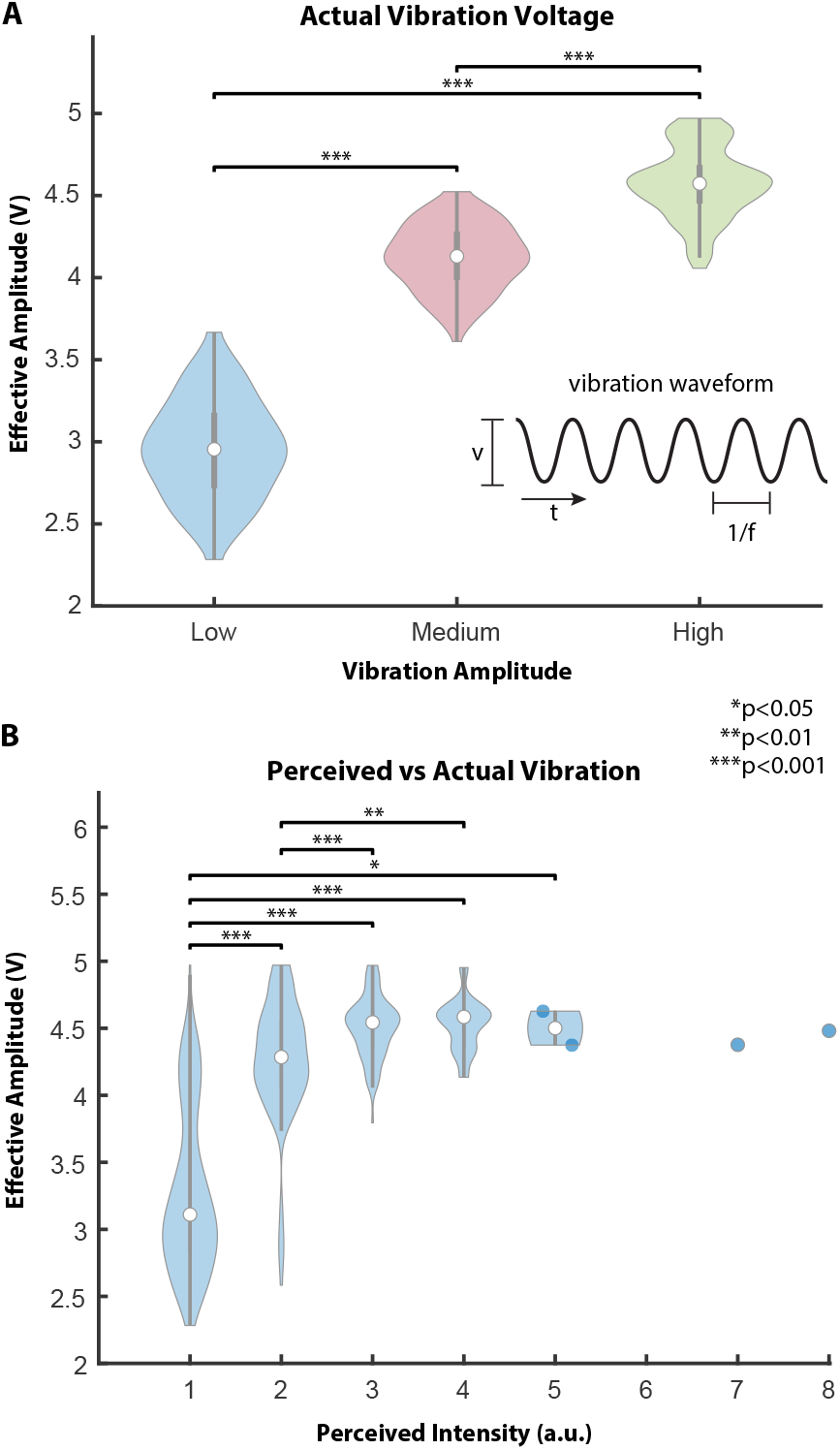
(A) The effective vibration amplitude had some variation, as measured by the voltage of the stimulation waveform, within a given amplitude condition. The vibration waveform duration (*t* = 500 ms) and frequency (*f* = 300 Hz) were set as constants in addition to the amplitude (i.e., voltage). (B) Effective vibration voltage as a function of subjective intensity ratings.

The subjective reported intensity increased with increasing vibration amplitude (Fig. 4B). It is not surprising that a larger vibration amplitude results in a stronger, more intense, haptic experience for the user. By matching reported intensity with the actual vibration amplitude voltage on each trial, it is clear that vibration amplitude alone is not necessarily indicative of the resulting perceived intensity at higher amplitude levels of stimulation. Specifically, the median tactor amplitude voltage on trials that were perceived as having an intensity rating of one and two were 3.1 V and 4.3 V, respectively; however, trials with perceived intensities of three through eight had a range of tactor amplitude voltages of only 4.49-4.59 V with no significant difference between any of those distributions (*p >* 0.05). Although there were only four trials that resulted in a perceived intensity rating of five or higher, there were 109 trials that elicited an intensity rating of either a three or four.

Interestingly, there were more than twice as many discrete values in the subjective reported intensities compared to the number of distinct vibration amplitude categories (low, medium, and high). This difference in the number of discrete values between the perceptual reports and the physical vibration indicates a finer perceptual resolution within the three discrete stimulation categories.

### E. High-gamma activity better reflects perceived intensity

To better understand how HG neural activity represents stimulation amplitude and the resulting perceived intensity, we performed a regression analysis on the data. We evaluated the relationship between HG spectral power and both the vibration amplitude and the perceived intensity by selecting the top five channels with the highest HG spectral power for each condition.

The linear regression model of HG spectral power and vibration amplitude (Fig. 5A) was statistically significant, *F* (1, 73) = 4.661*, p* = 0.0341, and explained approximately 6% of the variance (*R*^2^ = 0.060). The linear regression model of HG spectral power and perceived intensity (Fig. 5B) was statistically significant, *F* (1, 118) = 51.15*, p <* 0.0001, and explained approximately 30.2% of the variance (*R*^2^ = 0.302). Given the unique neural responses and activation level corresponding to specific vibration location (as shown in Fig. 1), regression analysis on each vibration location were performed. HG spectral power corresponding to middle finger appeared to have strongest relationship with both the vibration amplitude and the perceived intensity, with statistically significant *F* (1, 73) = 31.55*, p <* 0.0001 with *R*^2^ = 0.708 and *F* (1, 118) = 23.51*, p <* 0.0001 with *R*^2^ = 0.464, respectively.

**Fig. 5.**
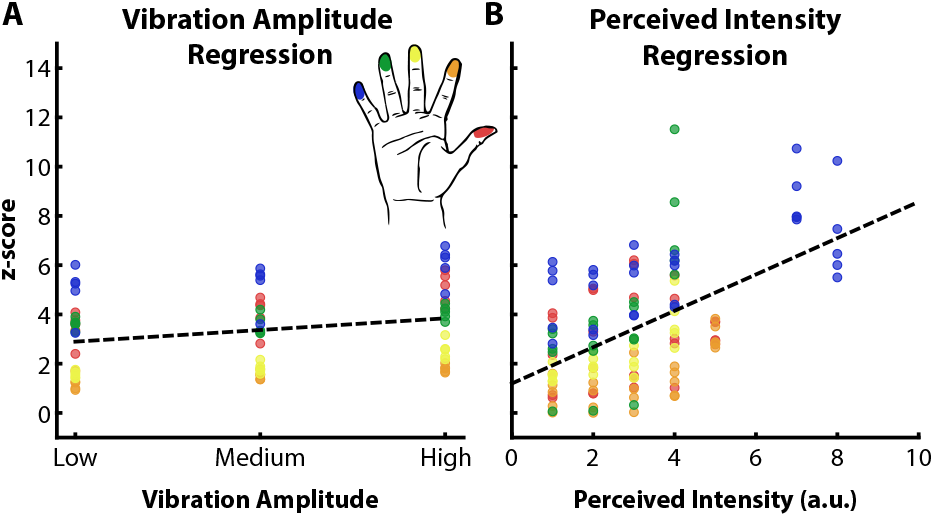
Linear regression model relating HG activity of the top five channels on each finger to (A) vibration amplitude and (B) perceived intensity of fingertip haptic stimulation. HG correlation with the perceived intensity was stronger than with vibration amplitude.

A Spearman’s rank-order was performed to assess the monotonic relationship between HG spectral power and both the vibration amplitude and the perceived intensity. There was weak association between HG spectral power and the vibration amplitude, with statistical significant positive correlation (*r_s_*(73) = 0.2799*, p* = 0.015). Whereas HG spectral power yielded moderate association with perceived intensity, with statistically significant positive correlation (*r_s_*(118) = 0.4479*, p <* 10^−6^).

These results show that HG power has a stronger relationship with the perceived intensity than the vibration amplitude.

## IV. Conclusion

In this study we demonstrated the feasibility of using neural activity as a way to identify perceived haptic intensity. Hand mapping identified unique cortical locations that corresponded to vibration at each individual fingertip, which revealed the localization of the neural response to the tactile stimulus. We found that HG activity in somatosensory cortex reflected both the vibration amplitude and perceived haptic intensity at different spectral power levels following the degree of vibration amplitude as well as perceived intensity. The stronger the perception, the higher the HG activation. Moreover, HG activity had stronger significant association with perceived intensity.

The findings revealed that perceived haptic intensity may not always align with physical stimulation amplitude, but HG activity could provide insights on the actual perceptual experience or quality, which could provide a useful signal for understanding how effective a haptic feedback strategy is for a BCI system.

## Data Availability

All data produced in the present work are contained in the manuscript

